# Investigating the validity of the Strengths and Difficulties Questionnaire to assess ADHD in young adulthood

**DOI:** 10.1101/2021.02.02.20248239

**Authors:** Lucy Riglin, Sharifah Shameem Agha, Olga Eyre, Rhys Bevan Jones, Robyn E Wootton, Ajay K Thapar, Stephan Collishaw, Evie Stergiakouli, Kate Langley, Anita Thapar

## Abstract

Attention Deficit Hyperactivity Disorder (ADHD) symptoms typically onset early in development and persist into adulthood for many. Robust investigation of symptom continuity and discontinuity requires repeated assessments using the same measure, but research is lacking into whether measures used to assess ADHD symptoms in childhood are also valid in adulthood. The Strengths and Difficulties Questionnaire (SDQ) has been widely used to measure ADHD symptoms in children, but little is known about its utility to measure ADHD in adulthood. We used Receiver Operating Characteristic (ROC) curve analyses to examine the validity of the SDQ hyperactivity/ADHD subscale to distinguish between cases and non-cases of DSM-5 ADHD classified using the Barkley Adult ADHD Rating Scale-IV (BAARS-IV) at age 25 years in a UK population cohort (N=4121). Analyses suggested that the SDQ ADHD subscale had high accuracy in distinguishing ADHD cases from non-cases in young adulthood (area under the curve=0.90, 95% CI=0.87-0.93) and indicated a lower cut-point for identifying those who may have an ADHD diagnosis in this age group compared to that currently recommended for younger ages.Our findings suggest that the SDQ is suitable for ADHD research across different developmental periods, which will aid the robust investigation of ADHD from childhood to young adulthood.

**Highlights:**

- The SDQ is widely used to measure ADHD symptoms in children
- We investigated the validity of the SDQ to assess ADHD at age 25 years
- The SDQ ADHD subscale had high accuracy in distinguishing DSM-5 ADHD cases from non-cases
- A lower cut-point is needed to identify ADHD diagnosis in young adulthood compared to younger ages
- The SDQ is appropriate for ADHD research across different development periods

## 1. Introduction

Attention Deficit Hyperactivity Disorder (ADHD) typically is a childhood-onset neurodevelopmental disorder characterized by symptoms of inattention and hyperactivity/impulsivity. Symptoms tend to decline across childhood and adolescence, but around 65% of individuals with a childhood diagnosis are estimated to show some level of symptom persistence into adulthood (Faraone et al., 2006) and estimates of the prevalence rate of adult ADHD are approximately 2.5% to 4.4% (Kessler et al., 2006; Simon et al., 2009). ADHD symptoms are associated with distress and impairment, including in educational, occupational, social and other settings, across childhood, adolescence and in adulthood (Asherson et al., 2012; Thapar and Cooper, 2016).

Given the life-course impacts of ADHD, there is a need to be able to assess symptoms across development to monitor and investigate correlates of symptom persistence and desistence. In particular, there is growing interest in the transition from childhood and adolescence to young adulthood (Ford, 2020) - a period that is associated both with changing demands and the transition from child/adolescent mental health services (CAMHS) to adult mental health services (AMHS). Robust investigation of symptom continuity and discontinuity requires repeated assessments using the same measure (Goodman et al., 2007); however, there is a lack of research into whether measures commonly used to assess ADHD symptoms in childhood and adolescence are also valid assessment tools in adulthood.

Adult ADHD has generally been assumed to be a continuation of childhood ADHD and this is supported by findings that adult ADHD symptoms and diagnosis show similar characteristics (e.g. high heritability, associations with other neurodevelopmental problems) when accounting for change in measure and informant (Larsson et al., 2014; Riglin et al., 2020; Rovira et al., 2020). However, the Diagnostic and Statistical Manual of Mental Disorders (5^th^ edition, DSM-5) criteria acknowledge some developmental differences by requiring fewer symptoms for a diagnosis of ADHD in adulthood compared to earlier in life (American Psychiatric Association, 2013). Research also suggests some change in the presentation of ADHD symptoms with age, whereby inattentive symptoms are more likely to persist whereas hyperactive-impulsive symptoms seem to become less common with age (Davidson, 2008; Willcutt et al., 2012). The validity of using child/adolescent based measures in adulthood cannot therefore be assumed.

The Strengths and Difficulties Questionnaire (SDQ) hyperactivity/ADHD subscale (Goodman, 1997) has been used widely to assess ADHD symptoms in children and adolescents in both research and clinical settings across different countries. Self, parent and teacher versions of the questionnaire have been validated for children and adolescents, with self-reports suitable for children aged 11 years or older (Goodman, 2001; Goodman et al., 2000; He et al., 2013). More recently an adult version of the SDQ has been developed (using almost identical wording for the ADHD subscale), which has been found to show similar psychometric properties to child/adolescent samples (Brann et al., 2018). However, to our knowledge the ADHD subscale of the SDQ has yet to be validated against ADHD diagnosis in adulthood.

The aim of this study was to examine the criterion validity of the SDQ as an assessment of ADHD symptoms in young adulthood, by examining its ability to discriminate DSM-5 ADHD cases from non-cases at age 25 years in a UK population cohort. We also explored whether a different cut-point to that recommended in childhood and adolescence is required to optimize sensitivity and specificity with respect to an adult diagnosis. Follow-up analyses included 1) separate analyses for males and females, and 2) assessing generalizability across subscales by comparing results for the ADHD subscale to the other SDQ (emotional, conduct, peer, prosocial) subscales.

## 2. Methods

### 2.1. Sample

We analyzed data from the Avon Longitudinal Study of Parents and Children (ALSPAC), a well-established prospective, longitudinal birth cohort study (Boyd et al., 2013; Fraser et al., 2013; Northstone et al., 2019) (total possible N=14,901). Details of this study are provided in the Supplementary Material. Ethical approval for the study was obtained from the ALSPAC Ethics and Law Committee and the Local Research Ethics Committees. Informed consent for the use of data collected was obtained from participants following the recommendations of the ALSPAC Ethics and Law Committee at the time. Individuals were included in our analyses when ADHD diagnosis data were available: primary analyses were conducted using self-reports as would typically be used in adult mental health services (N=4121), with secondary analyses conducted using parent-reports (N=4330) (see below).

### 2.2. Diagnoses

ADHD diagnosis was assessed at age 25 years using the self-rated Barkley Adult ADHD Rating Scale (BAARS-IV) (Barkley, 2011). The diagnostic coding was based on the criteria of the DSM-5 (American Psychiatric Association, 2013), and was constructed by SSA, KL and LR with advice from clinical psychiatrists OE and RBJ. The BAARS-IV includes the 18 DSM-5 ADHD items (range 0-18) and assesses current ADHD symptoms and ten domains of impairment. Each item was defined to be clinically significant if it was endorsed as occurring ‘often’ or ‘very often’ in-line with recommendations (Barkley, 2011). In line with DSM-5, diagnosis required the presence of five or more symptoms of inattention or hyperactivity/impulsivity, symptom onset to be prior to age 12 years (assessed retrospectively also using the BAARS-IV), and for current symptoms to be accompanied by impairment in social, academic, or occupational functioning (see Supplementary Table 1) (American Psychiatric Association, 2013).

**Table 1.** **Sensitivity and specificity for SDQ hyperactivity/ADHD cut-points compared against ADHD diagnosis**

| Cut-point | Sensitivity (95% CI) | Specificity (95% CI) |
| --- | --- | --- |
| ≥ 1 | 99.2% (95.5-100.0) | 11.9% (10.9-12.9) |
| ≥ 2 | 99.2% (95.5-100.0) | 26.7% (25.3-28.1) |
| ≥ 3 | 98.4% (94.2-99.8) | 43.5% (41.9-45.0) |
| ≥ 4 | 94.2% (88.4-97.6) | 61.9% (60.3-63.4) |
| ≥ 5 | 89.3% (82.3-94.2) | 76.7% (75.3-78.0) |
| ≥ 6 | 76.0% (67.4-83.3) | 88.7% (87.7-89.7) |
| ≥ 7 | 51.2% (42.0-60.4) | 94.6% (93.9-95.3) |
| ≥ 8 | 37.2% (28.6-46.4) | 98.2% (97.7-98.6) |
| ≥ 9 | 20.7% (13.8-29.0) | 99.6% (99.4-99.8) |
| ≥ 10 | 5.0% (1.84 –10.5) | 100.0% (99.8-100.0) |

### 2.3. Strengths and Difficulties Questionnaire (SDQ)

The five item self-rated version of the hyperactivity/ADHD subscale of the SDQ (Goodman, 1997) (range 0-10) was also completed at age 25 years. The ADHD subscale is well-validated in child and adolescent samples, for which the recommended cut-point for “high” symptoms (top 10% of the general population) is ≥7, with scores of 0-5 categorized as close to average (Green et al., 2005). In line with recommendations (www.sdqinfo.org) total ADHD subscale scores were derived using mean imputation for those with (≤2) of SDQ items missing. Follow-up analyses investigated the other SDQ subscales of emotional problems, conduct problems, peer problems and prosocial behavior (each subscale has five items, range 0-10).

### 2.4. Analyses

Data were analyzed using Stata version 14. Receiver Operating Characteristic (ROC) curve analyses were used to examine the ability of the SDQ to distinguish between cases and non-cases of ADHD classified using the BAARS-IV. A ROC curve was plotted (sensitivity vs 1-specificity) and the area under the curve (AUC) estimated. Stata’s *cvauroc* function (Luque-Fernandez et al., 2019) was used to minimize overfitting by implementing a 10-fold cross-validation and bootstrapping cross-validated AUC values to obtain bias-adjusted confidence intervals. The AUC assesses the diagnostic efficiency of the SDQ subscale in relation to meeting ADHD diagnostic criteria, whereby 0.5 indicates performance at chance level and 1.0 indicates perfect detection. Values < 0.7 are generally considered low and those >0.9 excellent (Swets, 1988).

Sensitivity reflects the ability to correctly identify all those with a diagnosis (true positives) whereas specificity reflects the ability to correctly identify all those without a diagnosis (true negatives). Optimum cut-points for ADHD diagnosis were selected as those that best balanced sensitivity and specificity, i.e. assuming that false positives and false negatives are equally undesirable, according to maximal Youden Index (sensitivity + specificity – 1) (Youden, 1950). Given that previous work recommending cut-points in child and adolescent samples were based on identifying the top 10% of the population (Goodman, 1997; Green et al., 2005), we also explored which cut-points would capture 10% of our sample. Positive predictive values (PPV) and negative predictive values (NPV) were calculated for proposed cut-points, which reflect the probability that those exceeding the cut-point have a diagnosis and the probability that those not meeting the cut-point do not have a diagnosis, respectively.

Follow-up analyses were conducted (i) separately for males and females, using Stata’s *roccomp* function (Cleves, 2002) to compare AUC values by sex, and (ii) using the other SDQ subscales to distinguish between cases and non-cases of ADHD. Secondary analyses were conducted using parent-reports instead of self-reports. As parents have been observed to be important informants for ADHD symptoms even in early adulthood (Barkley et al., 2002), we tested the validity of the parent-rated SDQ against parent-reported diagnosis (see Supplementary Material).

## 3. Results

### 3.1. ADHD symptom scores using SDQ

The overall mean self-rated SDQ hyperactivity/ADHD subscale score at age 25 years was 3.09 (SD=2.11) with a Cronbach’s alpha of 0.67, suggesting acceptable levels of internal consistency. As has been reported elsewhere (Riglin et al., 2020), the mean SDQ hyperactivity score was higher in males than in females (mean difference=0.36, 95% CI=0.22–0.50, p=2.3×10^−07^). Mean scores for those who met DSM-5 ADHD diagnostic criteria were 6.70 (SD=1.92) compared to 2.98 (SD=2.02) for those who did not meet diagnostic criteria (mean difference=3.72, 95% CI=3.36-4.08, p=3×10^−85^).

### 3.2. DSM-5 diagnosis of ADHD

Of those with available data, 2.9% met DSM-5 diagnostic criteria for ADHD at age 25 years (see Supplementary Table 1). At age 25 years, male sex was not associated with an increased likelihood of meeting ADHD criteria (3.0% in females and 2.9% in males: OR=0.97, 95% CI=0.66-1.43, p=0.89).

### 3.3. Receiver Operating Characteristic (ROC) curve analyses

ROC curve analyses suggested the SDQ ADHD subscale has high accuracy in distinguishing ADHD cases from non-cases (AUC=0.90, 95% CI=0.87-0.93), as shown in Figure 1. Sensitivity and specificity values for all possible SDQ ADHD subscale cut-points are shown in Table 1.

**Figure 1.**
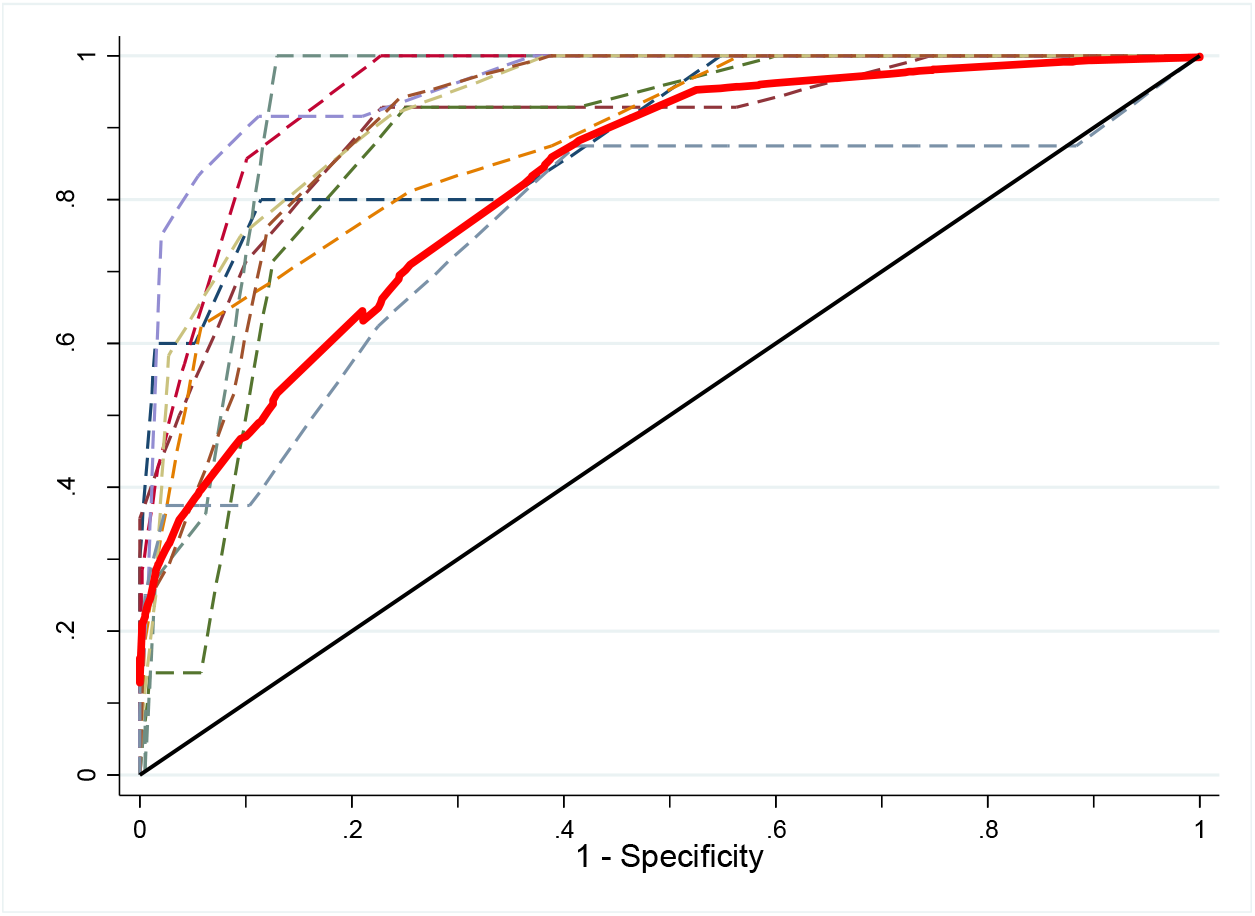
Receiver Operating Characteristic (ROC) curves for SDQ using ADHD diagnosis as criterion. Solid red curve = cross-validated AUC, dashed curves = k-fold ROC curves

### 3.4. Optimum cut-points

The optimum cut-point for distinguishing ADHD cases from non-cases was ≥5, which showed high accuracy in correctly identifying those meeting diagnostic criteria (i.e. true positives: sensitivity=89.3%) and good accuracy in correctly identifying those who did not meet diagnostic criteria (i.e. true negatives: specificity=76.7%). This cut-point captured 25.3% of the sample and was associated with a very high probability that those not meeting the cut-point did not meet diagnostic criteria (NPV=99.6%) but a lower probability that those exceeding the cut-point met diagnostic criteria (PPV=10.4%).

In line with previous work in younger samples (Goodman, 1997; Green et al., 2005) we also inspected the cut-points which identified the top 10% of the sample: this was ≥6 (sensitivity=76.0% and specificity=88.7%), which again was associated with a very high but slightly attenuated probability that those not meeting the cut-point did not meet diagnostic criteria (NPV=99.1%) but a slightly higher probability those exceeding the cut-point met diagnostic criteria (PPV=16.9%).

### 3.5. Analyses for males and females separately

When examining males and females separately, ROC curve analyses still indicated high accuracy in distinguishing ADHD cases from non-cases (males AUC=0.90, 95% CI=0.86-0.94; females AUC=0.90, 95% CI=0.87-0.94) with similar AUC values across sex (χ^2^(1)=0.02, p=0.89).

Sensitivity and specificity values for all possible SDQ subscale cut-points are shown in Table 2. The identified (whole-sample) cut-point of ≥5 captured 30.2% of males (NPV=99.6%, PPV=8.6%) and 22.7% of females (NPV=99.6%, PPV=11.6%) in our sample.

**Table 2.** **Sensitivity and specificity for SDQ hyperactivity/ADHD cut-points compared against ADHD diagnosis for males and females separately**

| Cut-point | Males |  | Females |  |
| --- | --- | --- | --- | --- |
|  | Sensitivity (95% CI) | Specificity (95% CI) | Sensitivity (95% CI) | Specificity (95% CI) |
| ≥ 1 | 100.0% (91.2-100.0) | 10.4% (8.8-12.2) | 98.8% (93.3-100.0) | 12.7% (11.4 – 14.0) |
| ≥ 2 | 100.0% (91.2-100.0) | 23.9% (21.6-26.2) | 98.8% (93.3-100.0) | 28.2% (26.4-29.9) |
| ≥ 3 | 100.0% (91.2-100.0) | 39.4% (36.8-42.0) | 97.5% (91.4-99.7) | 45.6% (43.6-47.5) |
| ≥ 4 | 97.5% (86.8-99.9) | 56.1% (53.4-58.8) | 92.6% (84.6-97.2) | 64.8% (63.0-66.6) |
| ≥ 5 | 90.0% (76.3-97.2) | 71.6% (69.1-73.9) | 88.9% (80.0-94.8) | 79.3% (79.7-80.8) |
| ≥ 6 | 77.5% (61.5-89.2) | 85.8% (83.8-87.6) | 75.3% (64.5-84.2) | 90.2% (89.0-91.3) |
| ≥ 7 | 57.5% (40.9-73.0) | 92.8% (91.3-94.1) | 48.2% (36.9-59.5) | 95.6% (94.7-96.3) |
| ≥ 8 | 35.0% (20.6-51.7) | 98.0% (97.1-98.7) | 38.3% (27.7-49.7) | 98.3% (97.7-98.8) |
| ≥ 9 | 20.0% (9.1-35.6) | 99.6% (99.0-99.8) | 21.0% (12.7-31.5) | 99.7% (99.4-99.8) |
| ≥ 10 | 5.0% (0.6-16.9) | 99.9% (99.6-100.0) | 4.9% (1.4 -12.2) | 100. 0% (99.8 -100.0) |

### 3.6. Other SDQ subscales

Analyses examining the other SDQ subscales found lower accuracy in distinguishing ADHD cases from non-cases; as shown in Table 3, AUC values ranged from 0.58 to 0.75 for the prosocial, conduct problems, peer problems and emotional problems subscales.

**Table 3.** **Area under the curve (AUC) for Strengths and Difficulties Questionnaire (SDQ) subscales compared against ADHD diagnosis**

|  | AUC (95% CI) |
| --- | --- |
| Hyperactivity/ADHD subscale | 0.90 (0.87-0.93) |
| Emotional problems subscale | 0.75 (0.70-0.79) |
| Conduct problems subscale | 0.65 (0.56-0.68) |
| Peer problems subscale | 0.72 (0.65-0.76) |
| Prosocial subscale | 0.58 (0.49-0.61) |
| Mean cross-validated AUC values and bootstrap bias corrected 95% confidence intervals |  |

### 3.7. Secondary analyses using parent-reports

Analyses using parent-reported data found that the parent-reported SDQ was also able to accurately identify ADHD diagnosis, (see Supplementary Material): AUC=0.97 (95% CI=0.92-0.98) but with a lower optimum cut-point of ≥4, capturing 13.2% of the sample.

## 4. Discussion

This study aimed to examine the validity of the SDQ as an assessment of ADHD symptoms at age 25 years in a UK population cohort. We found the hyperactivity/ADHD subscale to have high accuracy in distinguishing those meeting ADHD diagnostic criteria from those who did not. This suggests that the SDQ subscale, which is widely used in child and adolescent populations, is also suitable for use in young adults.

Our finding of excellent accuracy of the self-rated SDQ for measuring ADHD in adulthood (AUC=0.90) is similar if not somewhat higher than previous work in children and adolescents (using self- and parent-reports) (Algorta et al., 2016; Goodman et al., 2003), including previous work in the present sample at age 17 years (AUC=0.89) using parent-reports (Caye et al., 2019). In general, despite the trend that hyperactive-impulsive symptoms are likely to decline whilst inattentive symptoms persist with age (Davidson, 2008; Willcutt et al., 2012), the SDQ subscale - which includes three hyperactive-impulsive and two inattentive symptoms - shows high validity across childhood, adolescence and into adulthood. This suggests that while the presentation of symptoms may change with age, the SDQ items capture a similar construct. Indeed, previous work in a smaller high-risk sample found the SDQ to have similar psychometric properties (e.g. inter-scale correlations, internal consistency, and inter-rater agreement) in adults compared with adolescents (Brann et al., 2018).

We also investigated whether different cut-points are required to capture clinically relevant symptoms in young adulthood compared to younger ages. Current recommendations based on identifying the top 10% of the population suggest that in childhood and adolescence, self-rated scores of ≥7 capture those with high symptoms, with scores of 0-5 capturing those close to average (Goodman, 1997; Green et al., 2005). At age 25 years, our results suggest that lower cut-points are needed to capture clinically relevant symptoms, with scores of ≥5 achieving the best balance between sensitivity and specificity in identifying those meeting ADHD diagnostic criteria and a cut-point of ≥6 capturing 10% of our sample. These lower thresholds likely reflect the generally lower levels of ADHD symptoms in adulthood and is consistent with diagnostic criteria that require fewer symptoms to be present for an adult diagnosis to be made (American Psychiatric Association, 2013). While these cut-points performed well in identifying those meeting diagnostic criteria, they also captured a broader group of individuals, suggesting that this short measure is better used as a screening tool or to capture individuals with high levels of symptoms, rather than as a diagnostic tool. Replication of these cut-points in different samples is an important area for future research.

The selection of appropriate cut-points depends on the aim and rationale for using the instrument. We selected cut-points based on the premise that false positives and false negatives were equally undesirable (Youden, 1950), which is most likely appropriate for a one-stage approach in research settings to define a group most likely to have a diagnosis. However, for use in high-risk samples or as a clinical screening measure, possible cases would be followed up with further clinical assessment in a two-stage approach. In these circumstances, an increased number of false positives may be considered acceptable (Goodman, 1997) and lower cut-points that favor sensitivity may be preferable. In contrast, higher cut-points that favor specificity would be more appropriate for measures that are used for diagnostic purposes (Silva et al., 2015). The selection of cut points can also depend on sample characteristics such as age and sex. Our study suggests the need for a lower cut-point in young-adulthood to capture ADHD in young adulthood than in childhood/adolescence, but we found little evidence of sex differences in the ability of the SDQ to measure young adult ADHD, in line with previous work in children and adolescents (Algorta et al., 2016). Finally, we found the ADHD subscale of the SDQ to show higher accuracy in distinguishing between cases and non-cases of ADHD compared to the other SDQ subscales (emotional, conduct, peer, prosocial), which support the specificity of this subscale.

While our primary analyses focused on self-reports, we conducted secondary analyses using parent-reports (of both the SDQ and to define diagnosis). While adult mental health services may not typically involve parents, research studies often utilize parent ratings at earlier ages, making the validity of the parent-rated SDQ to assess ADHD in young adulthood important for researchers interested in assessing continuity and discontinuity, which ideally requires repeated assessments using the same measure and informant (Goodman et al., 2007). We found parent-ratings using the young adult SDQ to also show high accuracy and – consistent with our primary analyses using self-reports – results suggested that lower cut-points are needed to capture clinically relevant symptoms (≥4 in young adulthood compared to ≥8 in childhood/adolescence.)

The validity of parent ratings of their young adult offspring’s ADHD symptoms is consistent with recent work in this sample which found parent SDQ ratings of ADHD show similar genetic and neurodevelopmental correlates at age 25 as in childhood, suggesting that these reports capture a similar construct across development (Riglin et al., 2020) and implying that parents may still provide valuable insight into their offspring’s symptoms at 25 years.

Our study has a number of strengths, including the use of a large population sample and the investigation of a measure widely used in childhood and adolescence, which will enable continuity in future investigation of ADHD across different developmental periods. However, findings should also be considered in light of limitations, including non-random attrition which means those included in this sample will not be fully representative of the broader population, as those with elevated risk of psychopathology are more likely to have dropped out of this birth-cohort by the time data were collected at age 25 years (Martin et al., 2016; Taylor et al., 2018). This is particularly relevant to sample specific cut-points (i.e. capturing the top 10%) which will likely be lower than would be found in the general population. We were also reliant on questionnaire rather than direct interview data to generate ADHD diagnoses, although this is a widely used tool which covers all diagnostic criteria, including impairment and age-at-onset (Barkley, 2011). Finally, it is not clear the extent to which findings from this community sample are applicable to high-risk or clinical samples.

In conclusion, we found the widely-used and freely-available SDQ, used in children and adolescents, to be a valid measure for assessing ADHD symptoms in young adults. We identified a different, lower cut-point to help more accurately identify those who may have an ADHD diagnosis in this age group. Our findings suggest that the SDQ is suitable for ADHD research across different developmental periods, which will aid the robust investigation of ADHD from childhood to young adulthood.

## Data Availability

The study website contains details of all the data that is available through a fully searchable data dictionary and variable search tool: http://www.bristol.ac.uk/alspac/researchers/our-data/.

## Acknowledgement

We are extremely grateful to all the families who took part in this study, the midwives for their help in recruiting them, and the whole ALSPAC team, which includes interviewers, computer and laboratory technicians, clerical workers, research scientists, volunteers, managers, receptionists and nurses. The UK Medical Research Council and Wellcome (Grant ref: 217065/Z/19/Z) and the University of Bristol provide core support for ALSPAC. This publication is the work of the authors and Lucy Riglin, Sharifah Shameem Agha and Anita Thapar will serve as guarantors for the contents of this paper. A comprehensive list of grants funding is available on the ALSPAC website (www.bristol.ac.uk/alspac/external/documents/grant-acknowledgements.pdf). The measures used in the paper were specifically funded by the Wellcome Trust (204895/Z/16/Z). REW and ES work in a unit that receives funding from the University of Bristol and the UK Medical Research Council (MC_UU_00011/1 and MC_UU_00011/3). RBJ is supported by the Welsh Government through Health and Care Research Wales (National Institute for Health Research Fellowship, NIHR-PDF-2018). REW is supported by a postdoctoral fellowship from the South-Eastern Regional Health Authority (2020024). This study was supported by the Wellcome Trust (204895/Z/16/Z).

## Author contributions

LR, SSA, OE, RBJ and AT conceptualised the study, LR, SSA, OE, RBJ and KL derived the variables, LR wrote the original draft, SSA performed statistical analyses, ES and AT attained the study funding. All authors contributed to the interpretation of data and reviewed/edited the manuscript.

## References

Algorta, G.P., Dodd, A.L., Stringaris, A., Youngstrom, E.A., 2016. Diagnostic efficiency of the SDQ for parents to identify ADHD in the UK: a ROC analysis. Eur Child Adolesc Psychiatry 25(9), 949–957.

American Psychiatric Association, 2013. Diagnostic and statistical manual of mental disorders, 5th edition, 5th ed. American Psychiatric Association., Washington, D.C.

Asherson, P., Akehurst, R., Kooij, J.S., Huss, M., Beusterien, K., Sasané, R., Gholizadeh, S., Hodgkins, P., 2012. Under diagnosis of adult ADHD: cultural influences and societal burden. Journal of Attention Disorders 16(5_suppl), 20S–38S.

Barkley, R.A., 2011. Barkley Adult ADHD Rating Scale-IV (BAARS-IV). Guilford Press.

Barkley, R.A., Fischer, M., Smallish, L., Fletcher, K., 2002. The persistence of attention-deficit/hyperactivity disorder into young adulthood as a function of reporting source and definition of disorder. J Abnorm Psychol 111(2), 279–289.

Boyd, A., Golding, J., Macleod, J., Lawlor, D.A., Fraser, A., Henderson, J., Molloy, L., Ness, A., Ring, S., Davey Smith, G., 2013. Cohort Profile: the ‘children of the 90s’--the index offspring of the Avon Longitudinal Study of Parents and Children. International journal of epidemiology 42(1), 111–127.

Brann, P., Lethbridge, M.J., Mildred, H., 2018. The young adult Strengths and Difficulties Questionnaire (SDQ) in routine clinical practice. Psychiatry Res 264, 340–345.

Caye, A., Agnew-Blais, J., Arseneault, L., Gonçalves, H., Kieling, C., Langley, K., Menezes, A.M.B., Moffitt, T.E., Passos, I.C., Rocha, T.B., Sibley, M.H., Swanson, J.M., Thapar, A., Wehrmeister, F., Rohde, L.A., 2019. A risk calculator to predict adult attention-deficit/hyperactivity disorder: generation and external validation in three birth cohorts and one clinical sample. Epidemiol Psychiatr Sci 29, e37.

Cleves, M.A., 2002. From the Help Desk: Comparing Areas under Receiver Operating Characteristic Curves from Two or more Probit or Logit Models. The Stata Journal 2(3), 301–313.

Davidson, M.A., 2008. Literature Review: ADHD in Adults:A Review of the Literature. Journal of Attention Disorders 11(6), 628–641.

Faraone, S.V., Biederman, J., Mick, E., 2006. The age-dependent decline of attention deficit hyperactivity disorder: a meta-analysis of follow-up studies. Psychol Med 36(2), 159–165.

Ford, T., 2020. Transitional care for young adults with ADHD: transforming potential upheaval into smooth progression. Epidemiology and Psychiatric Sciences 29, e87.

Fraser, A., Macdonald-Wallis, C., Tilling, K., Boyd, A., Golding, J., Davey Smith, G., Henderson, J., Macleod, J., Molloy, L., Ness, A., Ring, S., Nelson, S.M., Lawlor, D.A., 2013. Cohort Profile: the Avon Longitudinal Study of Parents and Children: ALSPAC mothers cohort. International journal of epidemiology 42(1), 97–110.

Goodman, R., 1997. The Strengths and Difficulties Questionnaire: a research note. J Child Psychol Psychiatry 38(5), 581–586.

Goodman, R., 2001. Psychometric properties of the strengths and difficulties questionnaire. J Am Acad Child Adolesc Psychiatry 40(11), 1337–1345.

Goodman, R., Ford, T., Simmons, H., Gatward, R., Meltzer, H., 2000. Using the Strengths and Difficulties Questionnaire (SDQ) to screen for child psychiatric disorders in a community sample. Br J Psychiatry 177, 534–539.

Goodman, R., Iervolino, A.C., Collishaw, S., Pickles, A., Maughan, B., 2007. Seemingly minor changes to a questionnaire can make a big difference to mean scores: a cautionary tale. Soc Psychiatry Psychiatr Epidemiol 42(4), 322–327.

Goodman, R., Meltzer, H., Bailey, V., 2003. The Strengths and Difficulties Questionnaire: a pilot study on the validity of the self-report version. International review of psychiatry 15(1-2), 173–177.

Green, H., McGinnity, Á., Meltzer, H., Ford, T., Goodman, R., 2005. Mental health of children and young people in Great Britain, 2004. Palgrave Macmillan Basingstoke.

He, J.-P., Burstein, M., Schmitz, A., Merikangas, K.R., 2013. The Strengths and Difficulties Questionnaire (SDQ): the Factor Structure and Scale Validation in U.S. Adolescents. J Abnorm Child Psychol 41(4), 583–595.

Kessler, R.C., Adler, L., Barkley, R., Biederman, J., Conners, C.K., Demler, O., Faraone, S.V., Greenhill, L.L., Howes, M.J., Secnik, K., Spencer, T., Ustun, T.B., Walters, E.E., Zaslavsky, A.M., 2006. The prevalence and correlates of adult ADHD in the United States: results from the National Comorbidity Survey Replication. Am J Psychiatry 163(4), 716–723.

Larsson, H., Chang, Z., D’Onofrio, B.M., Lichtenstein, P., 2014. The heritability of clinically diagnosed attention deficit hyperactivity disorder across the lifespan. Psychol Med 44(10), 2223–2229.

Luque-Fernandez, M.A., Redondo-Sánchez, D., Maringe, C., 2019. cvauroc: Command to compute cross-validated area under the curve for ROC analysis after predictive modeling for binary outcomes. The Stata Journal 19(3), 615–625.

Martin, J., Tilling, K., Hubbard, L., Stergiakouli, E., Thapar, A., Davey Smith, G., O’Donovan, M.C., Zammit, S., 2016. Association of Genetic Risk for Schizophrenia With Nonparticipation Over Time in a Population-Based Cohort Study. Am J Epidemiol 183(12), 1149–1158.

Northstone, K., Lewcock, M., Groom, A., Boyd, A., Macleod, J., Timpson, N., Wells, N., 2019. The Avon Longitudinal Study of Parents and Children (ALSPAC): an update on the enrolled sample of index children in 2019. Wellcome Open Res 4, 51–51.

Riglin, L., Leppert, B., Langley, K., Thapar, A.K., O’Donovan, M.C., Davey Smith, G., Stergiakouli, E., Tilling, K., Thapar, A., 2020. Investigating attention-deficit hyperactivity disorder and autism spectrum disorder traits in the general population: What happens in adult life? Journal of Child Psychology and Psychiatry doi:10.1111/jcpp.13297.

Rovira, P., Demontis, D., Sánchez-Mora, C., Zayats, T., Klein, M., Mota, N.R., Weber, H., Garcia-Martínez, I., Pagerols, M., Vilar-Ribó, L., Arribas, L., Richarte, V., Corrales, M., Fadeuilhe, C., Bosch, R., Martin, G.E., Almos, P., Doyle, A.E., Grevet, E.H., Grimm, O., Halmøy, A., Hoogman, M., Hutz, M., Jacob, C.P., Kittel-Schneider, S., Knappskog, P.M., Lundervold, A.J., Rivero, O., Rovaris, D.L., Salatino-Oliveira, A., da Silva, B.S., Svirin, E., Sprooten, E., Strekalova, T., Arias-Vasquez, A., Sonuga-Barke, E.J.S., Asherson, P., Bau, C.H.D., Buitelaar, J.K., Cormand, B., Faraone, S.V., Haavik, J., Johansson, S.E., Kuntsi, J., Larsson, H., Lesch, K.-P., Reif, A., Rohde, L.A., Casas, M., Børglum, A.D., Franke, B., Ramos-Quiroga, J.A., Soler Artigas, M., Ribasés, M., Consortium, A.W.G.o.t.P.G., andMe Research, t., 2020. Shared genetic background between children and adults with attention deficit/hyperactivity disorder. Neuropsychopharmacology.

Silva, T.B.F., Osório, F.L., Loureiro, S.R., 2015. SDQ: discriminative validity and diagnostic potential. Front Psychol 6(811).

Simon, V., Czobor, P., Balint, S., Meszaros, A., Bitter, I., 2009. Prevalence and correlates of adult attention-deficit hyperactivity disorder: meta-analysis. Br J Psychiatry 194(3), 204–211.

Swets, J.A., 1988. Measuring the accuracy of diagnostic systems. Science 240(4857), 1285–1293.

Taylor, A.E., Jones, H.J., Sallis, H., Euesden, J., Stergiakouli, E., Davies, N.M., Zammit, S., Lawlor, D.A., Munafo, M.R., Davey Smith, G., Tilling, K., 2018. Exploring the association of genetic factors with participation in the Avon Longitudinal Study of Parents and Children. International journal of epidemiology 47(4), 1207–1216.

Thapar, A., Cooper, M., 2016. Attention deficit hyperactivity disorder. Lancet 387(10024), 1240–1250.

Willcutt, E.G., Nigg, J.T., Pennington, B.F., Solanto, M.V., Rohde, L.A., Tannock, R., Loo, S.K., Carlson, C.L., McBurnett, K., Lahey, B.B., 2012. Validity of DSM-IV attention deficit/hyperactivity disorder symptom dimensions and subtypes. Journal of abnormal psychology 121(4), 991–1010.

Youden, W.J., 1950. Index for rating diagnostic tests. Cancer 3(1), 32–35.

