## Supplemental information for "Investigating the validity of the Strengths and Difficulties Questionnaire to assess ADHD in young adulthood"

### Supplementary Material

#### The Avon Longitudinal Study of Parents and Children (ALSPAC)

Pregnant women resident in Avon, UK with expected dates of delivery 1st April 1991 to 31st December 1992 were invited to take part in the study. The initial number of pregnancies enrolled is 14,541 (for these at least one questionnaire has been returned or a "Children in Focus" clinic had been attended by 19/07/99). Of these initial pregnancies, there was a total of 14,676 foetuses, resulting in 14,062 live births and 13,988 children who were alive at 1 year of age. When the oldest children were approximately 7 years of age, an attempt was made to bolster the initial sample with eligible cases who had failed to join the study originally. As a result, the total sample size for data collected after the age of seven is therefore 15,454 pregnancies, resulting in 15,589 foetuses. Of these 14,901 were alive at 1 year of age. Part of this data was collected using REDCap (Harris et al., 2019; Harris et al., 2009). Please note that the study website contains details of all the data that is available through a fully searchable data dictionary and variable search tool: http://www.bristol.ac.uk/alspac/researchers/our-data/. Further details of the study, measures and sample can be found elsewhere (Boyd et al., 2013; Fraser et al., 2013; Northstone et al., 2019). Where families included multiple births, we included the oldest sibling.

#### Parent-reports

Secondary analyses were conducted using parent-reports. While adult mental health services may not typically involve parents, the validity of the parent-rated Strengths and Difficulties Questionnaire (SDQ) to assess ADHD is of importance for researchers interested in assessing continuity and discontinuity requires repeated assessments of the same measure (Goodman, Iervolino, Collishaw, Pickles, & Maughan, 2007).

##### *Measures*

ADHD diagnosis was assessed at age at 25 years using the same diagnostic coding used for self-reports -the parent-rated Barkley Adult ADHD Rating Scale (BAARS-IV) (Barkley, 2011) (see Supplementary Table 1).

The five item parent-rated version of the hyperactivity/ADHD subscale of the SDQ (Goodman, 1997) (range 0-10) was also completed at age 25 years, for which the recommended cut-point for "high" symptoms (top 10% of the general population) is ≥8 for parent-ratings, with scores of 0-5 categorized as close to average (Green, McGinnity, Meltzer, Ford, & Goodman, 2005). In line with recommendations (www.sdqinfo.org) total ADHD subscale scores were derived using mean imputation for those with (≤2) of SDQ items missing. Sensitivity analyses investigated the other SDQ subscales of emotional problems, conduct problems, peer problems and prosocial behavior (all five items, range 0-10).

***Results***

The overall mean parent-rated SDQ hyperactivity/ADHD subscale score was 1.54 (SD=1.8), Cronbach's alpha = 0.72, with higher mean scores observed for males compared to females (mean difference=0.34, 95% CI=0.23-0.45, $p=7.4 \times 10^{-10}$). Mean scores were higher for those who met ADHD diagnostic criteria compared to those who did not (6.98, SD=2.20 compared to 1.47, SD=1.69; mean difference=5.51, 95% CI=5.07-5.95, $p=1 \times 10^{-124}$).

Of those with available data, 1.3% met DSM-5 diagnostic criteria for ADHD at age 25 years according to parent-report (see Supplementary Table 1). Male sex was associated with an increased likelihood of meeting ADHD criteria according to parent-report (0.8% in females and 1.9% in males: OR=2.38, 95% CI 1.39-4.07, $p=2 \times 10^{-03}$). Of those with both self-rated and parent-rated data (N=2632), there was only fair informant agreement (Kappa = 0.23), with 0.3% of this sample meeting diagnostic criteria according to both raters, 1.8% for self-report only and 0.4% for parent-report only.

ROC curve analyses suggested the parent-reported SDQ subscale to have high accuracy in distinguishing ADHD cases from non-cases (AUC=0.97, 95% CI=0.92-0.98). Sensitivity and specificity values for all possible SDQ subscale cut-points are shown in Supplementary Table 2. The optimum cut-points for distinguishing ADHD cases from non-cases was ≥4 (sensitivity=94.8%, specificity 87.9%, NPV=99.9%, PPV=9.6%) which captured 13.2% of the sample and was the same cut-point which identified the top 10% - this therefore suggests a lower cut-point in adulthood compared to the recommended cut-point of ≥8 for parent-ratings in childhood and adolescence (Green et al., 2005).

Analyses stratified by sex found little evidence of differences for males and females (males AUC=0.98, 95% CI=0.96-0.99, females AUC=0.94, 95% CI=0.88-0.99, $\chi^2(1)=1.88$, p=0.17), with the identified cut-point of ≥4 identified capturing 16.3% of males (NPV=100.0%, PPV=11.8%) and 10.5% of females (NPV= 99.9%, PPV= 6.6%) in our sample.

Sensitivity analyses examining the other SDQ subscales found lower accuracy in distinguishing ADHD cases from non-cases compared to the ADHD subscale (emotional problems AUC=0.88,

95% CI=0.79-0.89; conduct problems AUC=0.87, 95% CI=0.77-0.89; peer problems AUC=0.88, 95% CI=0.76-0.90; prosocial AUC=0.86, 95% CI=0.77-0.89).

### References


Barkley, R. A. (2011). *Barkley Adult ADHD Rating Scale-IV (BAARS-IV)*: Guilford Press.

Boyd, A., Golding, J., Macleod, J., Lawlor, D. A., Fraser, A., Henderson, J., . . . Davey Smith, G. (2013). Cohort Profile: the 'children of the 90s'--the index offspring of the Avon Longitudinal Study of Parents and Children. *Int J Epidemiol, 42*(1), 111-127.

Fraser, A., Macdonald-Wallis, C., Tilling, K., Boyd, A., Golding, J., Davey Smith, G., . . . Lawlor, D. A. (2013). Cohort Profile: the Avon Longitudinal Study of Parents and Children: ALSPAC mothers cohort. *Int J Epidemiol, 42*(1), 97-110.

Goodman, R. (1997). The Strengths and Difficulties Questionnaire: a research note. *Journal of child psychology and psychiatry, and allied disciplines, 38*(5), 581-586.

Goodman, R., Iervolino, A. C., Collishaw, S., Pickles, A., & Maughan, B. (2007). Seemingly minor changes to a questionnaire can make a big difference to mean scores: a cautionary tale. *Social psychiatry and psychiatric epidemiology, 42*(4), 322-327.

Green, H., McGinnity, Á., Meltzer, H., Ford, T., & Goodman, R. (2005). *Mental health of children and young people in Great Britain, 2004*: Palgrave Macmillan Basingstoke.

Harris, P. A., Taylor, R., Minor, B. L., Elliott, V., Fernandez, M., O'Neal, L., . . . Duda, S. N. (2019). The REDCap consortium: Building an international community of software platform partners. *J Biomed Inform, 95*, 103208.

Harris, P. A., Taylor, R., Thielke, R., Payne, J., Gonzalez, N., & Conde, J. G. (2009). Research electronic data capture (REDCap)--a metadata-driven methodology and workflow process for providing translational research informatics support. *J Biomed Inform, 42*(2), 377-381.

Northstone, K., Lewcock, M., Groom, A., Boyd, A., Macleod, J., Timpson, N., & Wells, N. (2019). The Avon Longitudinal Study of Parents and Children (ALSPAC): an update on the enrolled sample of index children in 2019. *Wellcome open research, 4*, 51-51.

**Supplementary Table 1.** Generating ADHD diagnoses

| | Self-rated | Parent-rated |
|---|---|---|
| *Inattention symptoms* | | |
| 1. Fail to give close attention to details or make careless mistakes in my work | 252/4131 (6%) | 77/4551 (1.7%) |
| 2. Have difficulty sustaining my attention in tasks or fun activities | 404/4131 (10%) | 78/4559 (1.7%) |
| 3. Don't listen when spoken to directly | 184/4130 (5%) | 99/4571 (2%) |
| 4. Don't follow through on instructions and fail to finish work | 187/4122 (5%) | 92/4554 (2%) |
| 5. Have difficulty organising tasks and activities | 334/4126 (8%) | 145/4553 (3%) |
| 6. Avoid, dislike or reluctant to engage in work that requires sustained mental effort | 366/4125 (9%) | 135/4570 (3%) |
| 7. Lose things necessary for tasks or activities | 225/4127 (6%) | 149/4565 (3%) |
| 8. Easily distracted | 671/4133 (16%) | 165/4571 (3.6%) |
| 9. Forgetful in daily activities | 496/4129 (12%) | 136/4560 (3%) |
| *Hyperactive/impulsive symptoms* | | |
| 1. Fidget with hands or feet or squirm in seat | 579/4130 (14%) | 110/4571 (2%) |
| 2. Leave my seat in situations in which sitting is expected | 129/4130 (3%) | 42/4560 (0.9%) |
| 3. Feel restless | 622/4127 (15%) | 118/4567 (2.6%) |
| 4. Have difficulty engaging in leisure activities or doing fun things quietly | 248/4127 (6%) | 84/4561 (1.8%) |
| 5. Feel "on the go" or "driven by a motor" | 518/4123 (13%) | 246/4573 (5%) |
| 6. Talk excessively | 431/4131 (10%) | 185/4571 (4%) |
| 7. Blurt out answers before questions have been completed | 190/4133 (5%) | 73/4567 (1.6%) |
| 8. Have difficulty awaiting turn | 131/4133 (3%) | 56/4559 (1%) |
| 9. Interrupt or intrude on others | 157/4121 (3.8%) | 76/4535 (184%) |
| *Meet criteria for five (or more) inattention symptoms* | 219/4134 (5%) | 79/4572 (1.7%) |
| *Meet criteria for five (or more) hyperactive/impulsive symptoms* | 141/4133 (3%) | 42/4570 (0.9%) |
| ***ADHD symptom criteria met*** (inattentive and/or hyperactive/impulsive symptoms) | 263/4133 (6%) | 85/4567 (1.9%) |
| *Symptoms present prior to age 12 years* | 134/247[#] (54%) | 64/76* (84%) |
| *Clear evidence that the symptoms interfere with, or reduce the quality of, social, academic, or occupational functioning* | 240/262[##] (92%) | 80/84** (95%) |
| **Meets symptom, age of onset and impairment criteria** | 121/4122 (2.9%) | 61/4564 (1.3%) |

There are no questions on pervasiveness in the Barkley questionnaire. *[#] n=16 with missing age of onset data. [##] n=1 with missing impairment data. *n=9 with missing age of onset data. **n=1 with missing impairment data.*

**Supplementary Table 2 Sensitivity and specificity for SDQ hyperactivity/ADHD cut-points compared against ADHD diagnosis, based on parent-reports**

| | Whole sample | | Males | | Females | |
|---|---|---|---|---|---|---|
| Cut-point | Sensitivity | Specificity | Sensitivity | Specificity | Sensitivity | Specificity |
| ≥ 1 | 100.0% (93.8-100.0) | 37.7% (36.3-39.2) | 100.0% (91.0-100.0) | 35.9% (33.8-38.1) | 100.0% (82.4-100.0) | 39.2% (37.2-41.3) |
| ≥ 2 | 98.3% (90.8-100.0) | 62.2% (60.7-63.6) | 100.0% (91.0-100.0) | 59.1% (56.9-61.3) | 94.7% (74.0-99.9) | 64.8% (62.8-66.8) |
| ≥ 3 | 96.6% (88.1-99.6) | 77.6% (76.3-78.8) | 100.0% (91.0-100.0) | 74.3% (72.3-76.2) | 89.5% (66.9-98.7) | 80.4% (78.7-82.0) |
| ≥ 4 | 94.8% (85.6-98.9) | 87.9% (86.9-88.9) | 100.0% (91.0-100.0) | 85.3% (83.7-86.9) | 84.2% (60.4-96.6) | 90.1% (88.8-91.3) |
| ≥ 5 | 84.5% (72.6-92.7) | 93.2% (92.3-93.9) | 89.7% (75.8-97.1) | 91.6% (90.3-92.8) | 73.7% (48.8-90.9) | 94.5% (93.5-95.4) |
| ≥ 6 | 77.6% (64.7-87.5) | 96.6% (96.0-97.2) | 84.6% (69.5-94.1) | 95.7% (94.7-96.6) | 63.2% (38.4-83.7) | 97.4% (96.7-98.0) |
| ≥ 7 | 60.3% (46.6-73.0) | 98.5% (98.1-98.8) | 66.7% (49.8-80.9) | 97.7% (96.9 -98.3) | 47.4% (24.4-71.1) | 99.2% (98.8-99.5) |
| ≥ 8 | 43.1% (30.3-56.8) | 99.4% (99.2-99.6) | 51.2% (34.8-68.0) | 99.1% (98.6-99.5) | 26.3% (9.2-51.2) | 99.7% (99.4-99.9) |
| ≥ 9 | 27.6% (16.7-40.9) | 99.9% (99.7-99.9) | 30.8% (17.0-47.6) | 99.9% (99.6-100.0) | 21.1% (6.1-45.6) | 99.9% (99.6 -100.0) |
| ≥ 10 | 15.5% (7.4-27.4) | 100.0% (99.8-100.0) | 17.9% (7.5-33.5) | 100.0% (99.7-100.0) | 10.5% (1.3-33.1) | 100.0% (99.8-100.0) |
